# When mistrust in the government and scientists reinforce social inequalities in vaccination against Covid-19

**DOI:** 10.1101/2022.02.23.22271397

**Authors:** Nathalie Bajos, Alexis Spire, Léna Silberzan, Antoine Sireyjol, Florence Jusot, Laurence Meyer, Jeanna-Eve Franck, Josiane Warszawski, the EpiCov study group

## Abstract

**Objective:** To assess whether mistrust in the government and scientists reinforces social and racial inequalities in vaccination practises

**Design:** A follow-up of a random population-based cohort survey.

**Setting:** In July 2021, in France.

**Participants:** 80,971 participants aged 18 years and more.

**Main outcome measures:** Adjusted odds ratios of Covid-19 vaccination status (received at least one dose/ intends to get vaccinated/ does not know whether to get vaccinated/refuses vaccination) were assessed using multinomial regressions to test associations with social and mistrust factors and to study how these two factors interacted with each other.

**Results:** In all, 72.2% were vaccinated at the time of the survey. The population of unvaccinated people was younger, less educated, had lower incomes, and more often belonged to racialised minorities, as compared to vaccinated people. Mistrust of government and scientists to curb the spread of the epidemic were the factors most associated with refusing to be vaccinated: OR=8.86 (7.13 to 11.00) for the government and OR=9.07 (7.71 to 10.07) for scientists, compared to vaccinated people. Mistrust was more prevalent among the poorer which consequently reinforced social inequalities in vaccination. The 10% poorest who did not trust the government reached an OR of 16.2 (11.9 to 22) for refusing to be vaccinated compared to the 10% richest who did.

**Conclusion:** There is a need to develop depoliticised outreach programmes targeted at the most socially disadvantaged groups, and to design vaccination strategies conceived with people from different social and racial backgrounds to enable them to make fully informed choices.

## INTRODUCTION

Many authors have emphasised the fact that Covid-19 vaccination is effective in combating the spread of the epidemic, as well as in reducing social inequalities in morbidity and mortality, provided that access to the Covid-19 vaccines is free and easy.[1–3] Making Covid-19 vaccines available does not necessarily lead to a very large population vaccine coverage, as shown by the percentages of people who are still not vaccinated in Western countries [4], even when these vaccines are free. Recent studies in the UK, in the US and in Norway [5–10] have shown that the most socially disadvantaged and racialised social groups are the least vaccinated. In light of their high risk of infection and mortality from Covid-19 [1], it appears all the more important to understand why they are less likely to be reached by Covid-19 vaccine programmes.

Social barriers hampering access to preventive practices, such as social distance from health professionals, geographical distance from health centres, or experiences of discrimination in the health system [11,12] need to be taken into account to study this particular preventive practice, vaccination. But in a context where governments have taken the lead in managing the pandemic crisis, it is all the more important to analyse vaccination practices accompanied by consideration of the trust that people place in the government. Many studies have shown the implications of political mistrust in Covid-19 vaccination intentions [13,14], but only a few have analysed its implications in vaccination practises. These studies were conducted in the US and showed that in counties with a high percentage of Republican voters, vaccination rates were significantly lower [15,16]. However, they were conducted at county or state level, and did not account for individual social characteristics. Furthermore, focusing on votes excludes people who are over-represented in the lower socio-economic groups [17–19], who have no political opinion or refuse to express themselves through voting. One study explored the relationship between trust in the government and racial differences in vaccine uptake in the US [20]; the results showed that trust in the government’s response was not indicative of vaccination. In any case, there is a need to clarify whether, and to what extent, mistrust in the government has an impact on vaccination practises. Because underprivileged social groups are known to be particularly mistrustful of the government [21–23], it could be thought that the government’s strong involvement in vaccine programmes and its high degree of politicisation are not likely to reinforce social inequalities in vaccination. In France as in many countries, the government strongly relied on scientists to justify its epidemic response actions. Studying the impact of trust in the government on vaccination practices therefore also implies taking trust in scientists into account. The objective of this article was *(i)* to identify social differences in vaccination status and mistrust in the government and scientists *(ii)* to investigate whether mistrust in the government and scientists increases social inequalities in vaccination practises.

## PARTICIPANTS AND METHODS

The EpiCov study consisted of a random sample of people aged 15 and over, excluding those living in institutional settings, selected from the FIDELI tax registry of the National Institute of Statistics and Economic Studies (INSEE), which covers 96% of the population living in France. Among them, 134,391 individuals participated in the first wave of the EpiCov study in May 2020 by answering the questionnaire either online or by phone. Among them, 107,808 (80.2%) respondents participated in the second wave in November 2020, which included questions on attitudes toward vaccination and 85,032 (79.0% from the second wave and 63.3% from the first one) in the third wave conducted in July 2021, which included questions on vaccination practises, and served as the basis for this analysis. We focused on people living in metropolitan France and over 18 years of age since vaccination was allowed only for adults at the time of the survey. In all, 80,971 (95.2%) individuals were included in our study.

Data collected included socio-demographic characteristics, household size and composition, a detailed description of comorbidities, health care use for Covid-19 and other symptoms, employment characteristics, vaccination and attitudes toward vaccination and other individual prevention measures during outings (alcohol-based hand sanitiser, mask, social distancing).

The survey was approved by the CNIL (French independent administrative authority responsible for data protection) on April 25th 2020 (ref: MLD/MFI/AR205138) and by the “Comité de protection des personnes” (French equivalent of the Research Ethics Committee) on April 24th. The survey also obtained an agreement from the “Comité du Label de la statistique publique”, proving its consistency with statistical quality standards.

### Outcome measures

Vaccination status was classified into 4 categories: vaccinated (at least one dose); intends to be vaccinated; does not know whether to get vaccinated; refuses vaccination.

Vaccinated people were also asked to give the date of their first injection.

### Social variables

We considered the following variables: age, gender, ethno-racial status (based on migration history), having children, social class (based on current or most recent occupation), if the respondent was a healthcare professional, standard of living (based on decile of income per household consumption unit) and formal education (defined according to the French hierarchical grid of educational qualifications). The ethno-racial status, used for the first time in France in a Covid survey, distinguished the mainstream population, *i*.*e*., people residing in metropolitan France who are neither immigrants nor native to French Overseas Departments (FOD, *i*.*e*. Martinique, Guadeloupe, Reunion Island, Guyane and Mayotte), nor descendants of immigrant(s) or native to FOD. For the minority population, a distinction was made between first-generation (immigrants) and second-generation (descendants of immigrants) immigrants, and the country of origin. The term racialised refers to immigrants or descendants of immigrants from the Maghreb, Turkey, Asia and Africa.[24]

### Living condition variables

We took two variables into account: the household composition (Lives alone/Lives with partner only/Lives with people other than partner only), and the population size of the municipality (Rural area/<100,000 inhabitants/≥100,000 inhabitants/Paris area). An ecological indicator was used to assess whether individuals lived in an area recognised as requiring additional public resources due to its high level of deprivation, known as “priority neighbourhood”.

### Health variables

Health variables included the existence of Covid-19 comorbidities (*i*.*e*., asthma or other respiratory diseases, high blood pressure or cardiovascular diseases, diabetes, cancer, HIV, mental or psychiatric disability, or BMI≥30) and if the respondent had had a positive Covid-19 test in the past 6 months.

Moreover, two variables from the second wave of the survey (November 2020) were used to assess perceived health status (Very good/Good/Fair/Poor/Very poor) and the perception of the Covid-19 risk (Afraid of contracting the virus/Not afraid of contracting the virus/No chance of contracting the virus/Does not know).

### Trust variables

Specific interest was finally devoted to the level of trust in the government (“To curb the spread of the coronavirus, what is the level of trust you place in the actions undertaken by (i) the government and (ii) by scientists?”: Complete trust/Fair amount of trust/Little trust/No trust at all/You do not know).

### Other variables

Finally, knowing a relative who has had a severe form of Covid-19, and the date of response to the questionnaire were also added in the analyses.

### Statistical analyses

A first univariate analysis was performed, to compare the distribution of the four categories of vaccination status according to social characteristics and mistrust variables. Then, the cumulative monthly rates of vaccination (from January 31st to June 30th 2021) were stratified by vaccination age categories (18-54/55-74/75+), and assessed according to formal education, standard of living and ethno-racial status. This allowed us to quantify the gap and the delay in vaccination between the most and the least disadvantaged group of people.

A multinomial regression was developed to compare the vaccinated people to the others (intend to be vaccinated; do not know whether to get vaccinated; refuse vaccination) and to investigate how non-vaccinated people differed among themselves according to social and mistrust variables.

The association between trust variables and social characteristics in vaccination status was studied using variables combining these two parameters. We created six twelve-modality variables crossing a binary variable characterising the trust variable (Complete trust/Fair amount of trust *versus* Little trust/No trust at all/I do not know, labelled as Trust+/Trust-) and formal education or standard of living or ethno-racial status. Six multinomial regressions were then performed, each one adjusted for one combining variable at a time.

Final calibrated weights were calculated to correct for non-response, as detailed elsewhere[25] for the first, second and third waves of the EpiCov survey. Response homogeneity groups were derived from the sampling weight divided by the probability of response estimated with logit models adjusted for auxiliary variables potentially linked to both the response mechanism and the main variables of interest in the EpiCov survey (age, gender, French *départment*, educational level, and region). The percentages presented are weighted to account for the sampling design with unequal inclusion probabilities due to an oversampling of low income populations and correction of nonresponse bias.

A P-value <0.05 was considered statistically significant. Given the sample size, the observed differences were consistently statistically significant. Therefore, no tests are presented for univariable analyses.

Individuals who answered that they did not wish to respond to the question on their vaccination status and/or the date of their first injection were excluded (n=193, 0.2%). Missing data was rare for all variables (<4%) and was deleted in multivariate analyses (n=7068, 8.7% excluded).

Statistics were done using R 4.1.1 [26], the tidyverse [27], the gtsummary [28], the nnet [29], and the Ggally [30] packages.

## RESULTS

The distribution of the vaccination status in the population is presented in Table 1. In all, 72.2% were vaccinated, with at least one injection in July 2021 (71.1% for men *versus* 73.3% for women). Less than one respondent out of ten (8.1%) refused to get vaccinated (8.2% of men and 8.1% of women), while one in ten (9.8%) said they intended to get vaccinated (10.7% for men and 9% for women), and a similar proportion did not yet know whether or not they would get vaccinated (10% for men and 9.7% for women).

**Table 1:**
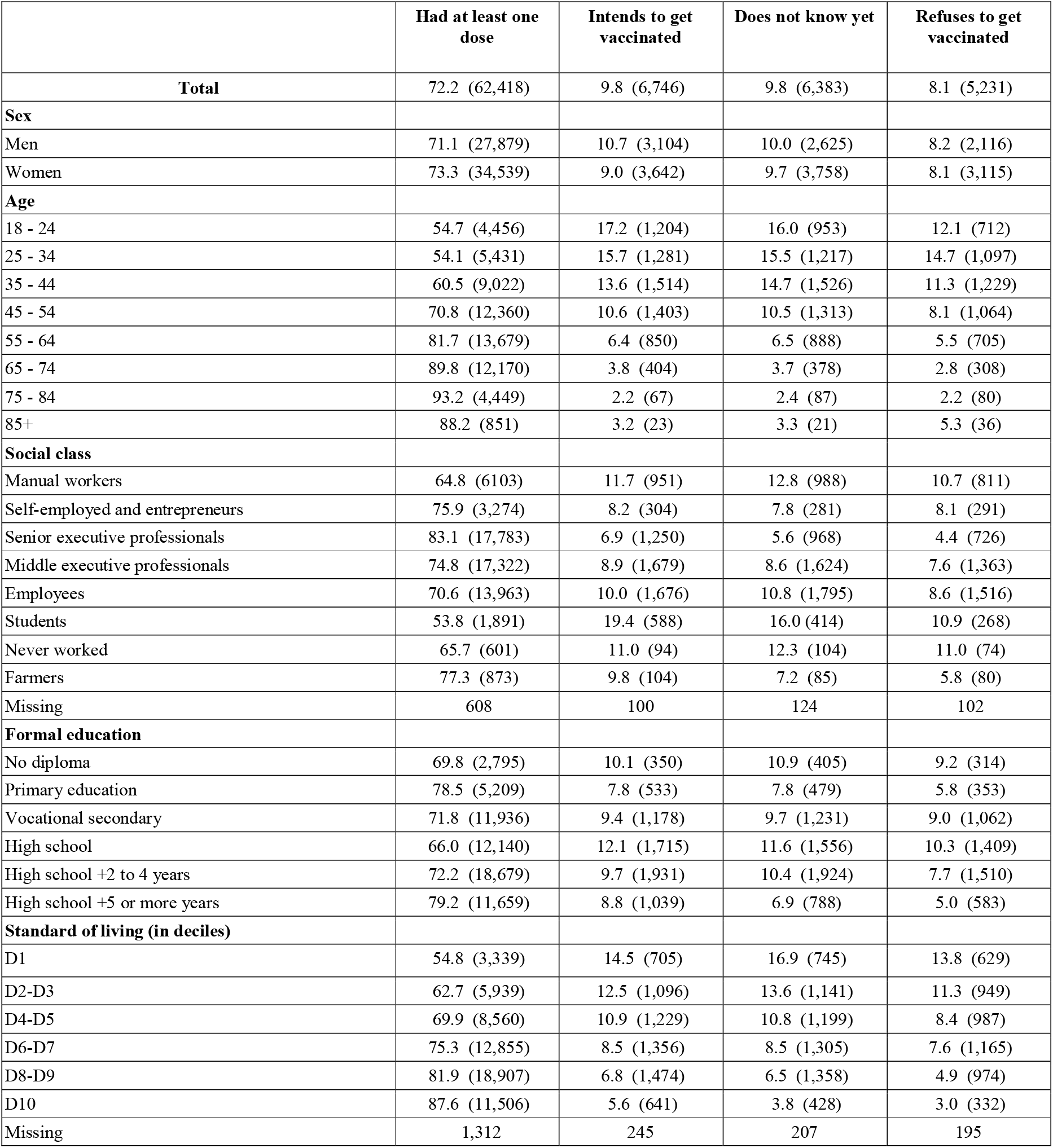
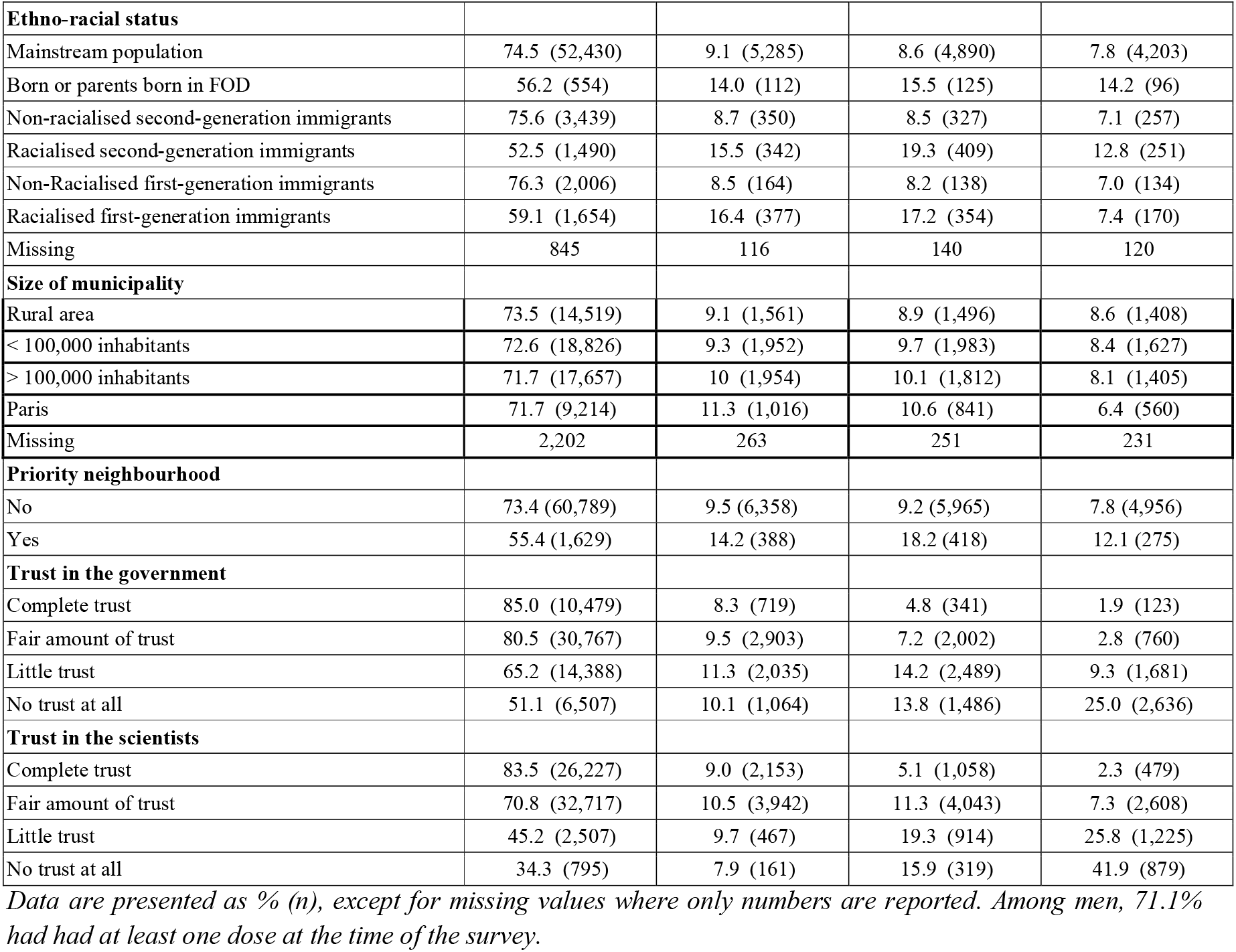
Distribution of the vaccinal status of people aged 18 years or over living in metropolitan France, by socio-demographic characteristics and mistrust variables. EpiCov study 3rd wave, July 2021.

|  | Had at least one dose | Intends to get vaccinated | Does not know yet | Refuses to get vaccinated |
| --- | --- | --- | --- | --- |
| **Total** | 72.2 (62,418) | 9.8 (6,746) | 9.8 (6,383) | 8.1 (5,231) |
| **Sex** |  |  |  |  |
| Men | 71.1 (27,879) | 10.7 (3,104) | 10.0 (2,625) | 8.2 (2,116) |
| Women | 73.3 (34,539) | 9.0 (3,642) | 9.7 (3,758) | 8.1 (3,115) |
| **Age** |  |  |  |  |
| 18 - 24 | 54.7 (4,456) | 17.2 (1,204) | 16.0 (953) | 12.1 (712) |
| 25 - 34 | 54.1 (5,431) | 15.7 (1,281) | 15.5 (1,217) | 14.7 (1,097) |
| 35 - 44 | 60.5 (9,022) | 13.6 (1,514) | 14.7 (1,526) | 11.3 (1,229) |
| 45 - 54 | 70.8 (12,360) | 10.6 (1,403) | 10.5 (1,313) | 8.1 (1,064) |
| 55 - 64 | 81.7 (13,679) | 6.4 (850) | 6.5 (888) | 5.5 (705) |
| 65 - 74 | 89.8 (12,170) | 3.8 (404) | 3.7 (378) | 2.8 (308) |
| 75 - 84 | 93.2 (4,449) | 2.2 (67) | 2.4 (87) | 2.2 (80) |
| 85+ | 88.2 (851) | 3.2 (23) | 3.3 (21) | 5.3 (36) |
| **Social class** |  |  |  |  |
| Manual workers | 64.8 (6103) | 11.7 (951) | 12.8 (988) | 10.7 (811) |
| Self-employed and entrepreneurs | 75.9 (3,274) | 8.2 (304) | 7.8 (281) | 8.1 (291) |
| Senior executive professionals | 83.1 (17,783) | 6.9 (1,250) | 5.6 (968) | 4.4 (726) |
| Middle executive professionals | 74.8 (17,322) | 8.9 (1,679) | 8.6 (1,624) | 7.6 (1,363) |
| Employees | 70.6 (13,963) | 10.0 (1,676) | 10.8 (1,795) | 8.6 (1,516) |
| Students | 53.8 (1,891) | 19.4 (588) | 16.0 (414) | 10.9 (268) |
| Never worked | 65.7 (601) | 11.0 (94) | 12.3 (104) | 11.0 (74) |
| Farmers | 77.3 (873) | 9.8 (104) | 7.2 (85) | 5.8 (80) |
| Missing | 608 | 100 | 124 | 102 |
| **Formal education** |  |  |  |  |
| No diploma | 69.8 (2,795) | 10.1 (350) | 10.9 (405) | 9.2 (314) |
| Primary education | 78.5 (5,209) | 7.8 (533) | 7.8 (479) | 5.8 (353) |
| Vocational secondary | 71.8 (11,936) | 9.4 (1,178) | 9.7 (1,231) | 9.0 (1,062) |
| High school | 66.0 (12,140) | 12.1 (1,715) | 11.6 (1,556) | 10.3 (1,409) |
| High school +2 to 4 years | 72.2 (18,679) | 9.7 (1,931) | 10.4 (1,924) | 7.7 (1,510) |
| High school +5 or more years | 79.2 (11,659) | 8.8 (1,039) | 6.9 (788) | 5.0 (583) |
| **Standard of living (in deciles)** |  |  |  |  |
| D1 | 54.8 (3,339) | 14.5 (705) | 16.9 (745) | 13.8 (629) |
| D2-D3 | 62.7 (5,939) | 12.5 (1,096) | 13.6 (1,141) | 11.3 (949) |
| D4-D5 | 69.9 (8,560) | 10.9 (1,229) | 10.8 (1,199) | 8.4 (987) |
| D6-D7 | 75.3 (12,855) | 8.5 (1,356) | 8.5 (1,305) | 7.6 (1,165) |
| D8-D9 | 81.9 (18,907) | 6.8 (1,474) | 6.5 (1,358) | 4.9 (974) |
| D10 | 87.6 (11,506) | 5.6 (641) | 3.8 (428) | 3.0 (332) |
| Missing | 1,312 | 245 | 207 | 195 |
| **Ethno-racial status** |  |  |  |  |
| Mainstream population | 74.5 (52,430) | 9.1 (5,285) | 8.6 (4,890) | 7.8 (4,203) |
| Born or parents born in FOD | 56.2 (554) | 14.0 (112) | 15.5 (125) | 14.2 (96) |
| Non-racialised second-generation immigrants | 75.6 (3,439) | 8.7 (350) | 8.5 (327) | 7.1 (257) |
| Racialised second-generation immigrants | 52.5 (1,490) | 15.5 (342) | 19.3 (409) | 12.8 (251) |
| Non-Racialised first-generation immigrants | 76.3 (2,006) | 8.5 (164) | 8.2 (138) | 7.0 (134) |
| Racialised first-generation immigrants | 59.1 (1,654) | 16.4 (377) | 17.2 (354) | 7.4 (170) |
| Missing | 845 | 116 | 140 | 120 |
| **Size of municipality** |  |  |  |  |
| Rural area | 73.5 (14,519) | 9.1 (1,561) | 8.9 (1,496) | 8.6 (1,408) |
| < 100,000 inhabitants | 72.6 (18,826) | 9.3 (1,952) | 9.7 (1,983) | 8.4 (1,627) |
| > 100,000 inhabitants | 71.7 (17,657) | 10 (1,954) | 10.1 (1,812) | 8.1 (1,405) |
| Paris | 71.7 (9,214) | 11.3 (1,016) | 10.6 (841) | 6.4 (560) |
| Missing | 2,202 | 263 | 251 | 231 |
| **Priority neighbourhood** |  |  |  |  |
| No | 73.4 (60,789) | 9.5 (6,358) | 9.2 (5,965) | 7.8 (4,956) |
| Yes | 55.4 (1,629) | 14.2 (388) | 18.2 (418) | 12.1 (275) |
| **Trust in the government** |  |  |  |  |
| Complete trust | 85.0 (10,479) | 8.3 (719) | 4.8 (341) | 1.9 (123) |
| Fair amount of trust | 80.5 (30,767) | 9.5 (2,903) | 7.2 (2,002) | 2.8 (760) |
| Little trust | 65.2 (14,388) | 11.3 (2,035) | 14.2 (2,489) | 9.3 (1,681) |
| No trust at all | 51.1 (6,507) | 10.1 (1,064) | 13.8 (1,486) | 25.0 (2,636) |
| **Trust in the scientists** |  |  |  |  |
| Complete trust | 83.5 (26,227) | 9.0 (2,153) | 5.1 (1,058) | 2.3 (479) |
| Fair amount of trust | 70.8 (32,717) | 10.5 (3,942) | 11.3 (4,043) | 7.3 (2,608) |
| Little trust | 45.2 (2,507) | 9.7 (467) | 19.3 (914) | 25.8 (1,225) |
| No trust at all | 34.3 (795) | 7.9 (161) | 15.9 (319) | 41.9 (879) |

*Data are presented as % (n), except for missing values where only numbers are reported. Among men, 71.1% had had at least one dose at the time of the survey.*

The vaccination rate increased very steadily with age, rising from 54.7% among 18-24 year olds to 93.2% among 75-84 year olds, and then falling to 88.2% among those over 85. There were also marked differences in vaccination practises according to social position. Only 69.8% of people without educational qualifications were vaccinated, compared to 79.2% of those with the highest qualifications. Similarly, 64.8% of manual workers had been vaccinated, compared to 83.1% of senior executives. As for the rate of vaccination according to income, it increased regularly from 54.8% among the 10% poorest to 87.6% among the 10% richest. Compared to the mainstream population (74.5%), the data showed that vaccination uptake was lower only among people belonging to racialised minorities, *i*.*e*. among first (59.1%) and second-generation racialised immigrants (52.5%) and among people born or whose parents were born in French Overseas Departments (56.2%). Living in a populated area was not associated with being vaccinated, although living in a “priority neighbourhood” was (55.4% vs 73.4%).

Social differences were also found among unvaccinated people: the 10% richest and those with the highest qualifications were more likely to intend to accept vaccination (5.6% and 8.8% respectively) whereas the 10% poorest and people without qualifications were more likely to hesitate (16.9% and 10.9% respectively). Interestingly, racialised first-generation immigrants were among those who least often refused vaccination (7.4%) whereas people from the overseas territories were the most reluctant (14.2%).

The data also showed that social differences were present even before vaccination and that they were maintained or widened over time, especially among the 18-54 year-olds (figure 1a, figure 1b, figure 1c). Among these, the gap between the 10% poorest and the 10% richest was 11.9 at the end of April 2021, a few days before vaccination was officially opened to everyone in this age group regardless of their comorbidities, and it increased to 35.4 by the end of June. In other words, the level of vaccination reached by the 10% poorest at the end of June 2021 had already been achieved by the 10% richest more than a month earlier.

**Figure 1a:**
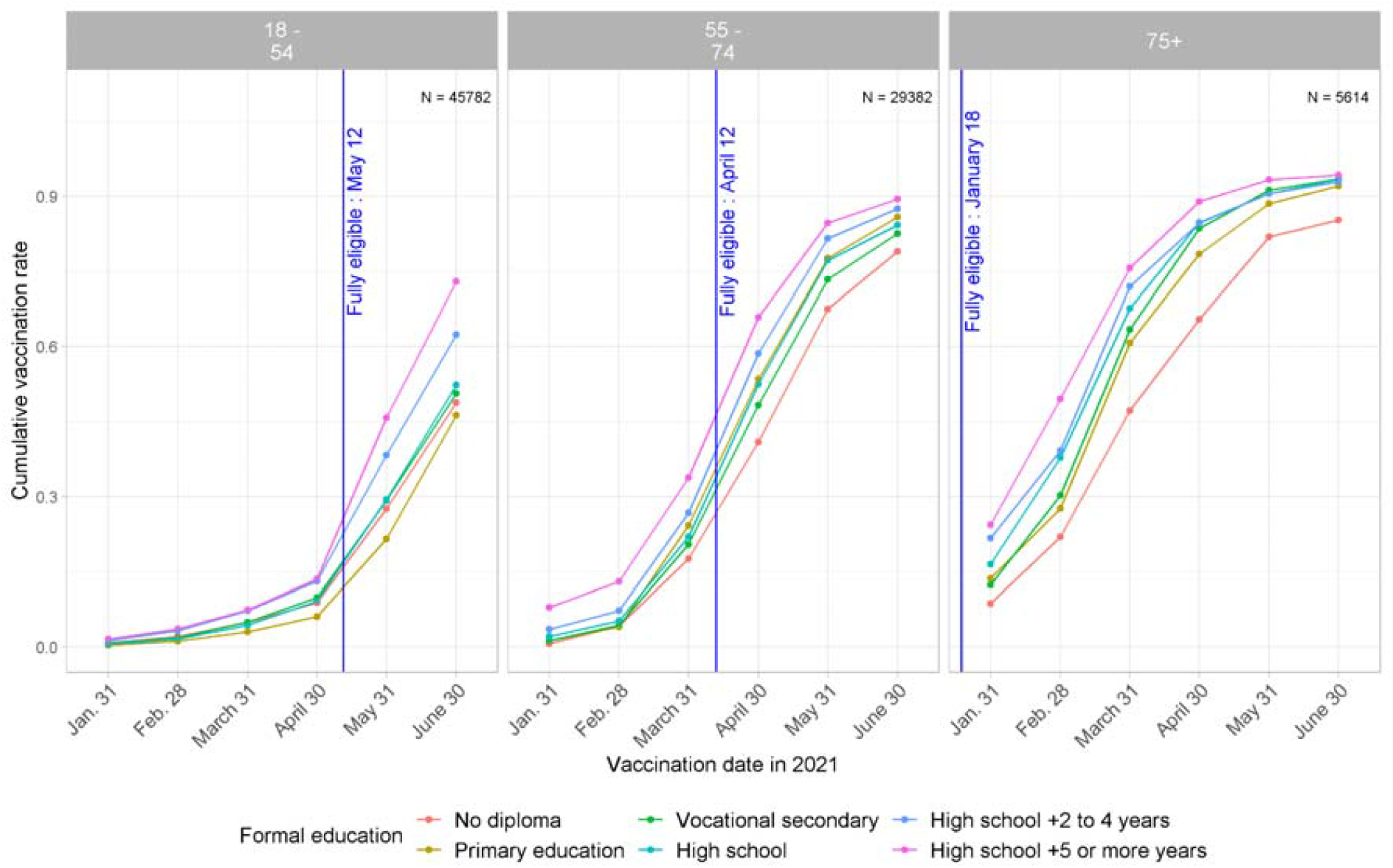
Trends over time in vaccination cumulative incidence rates by age, according to level of education. EpiCov study, 3rd wave, July 2021.

**Figure 1b:**
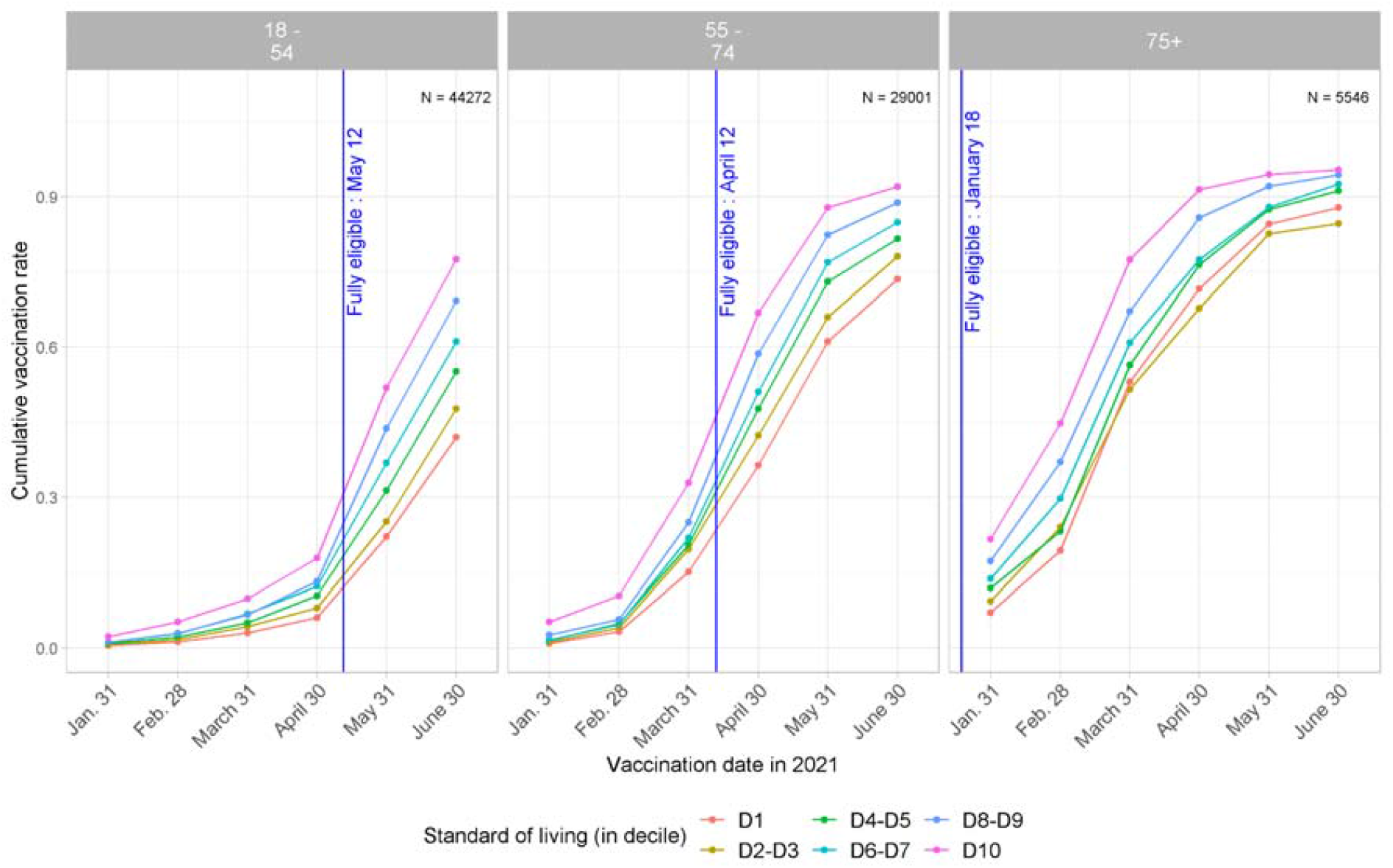
Trends over time in vaccination cumulative incidence rates by age, according standard of living (in decile). EpiCov study, 3rd wave, July 2021.

**Figure 1c:**
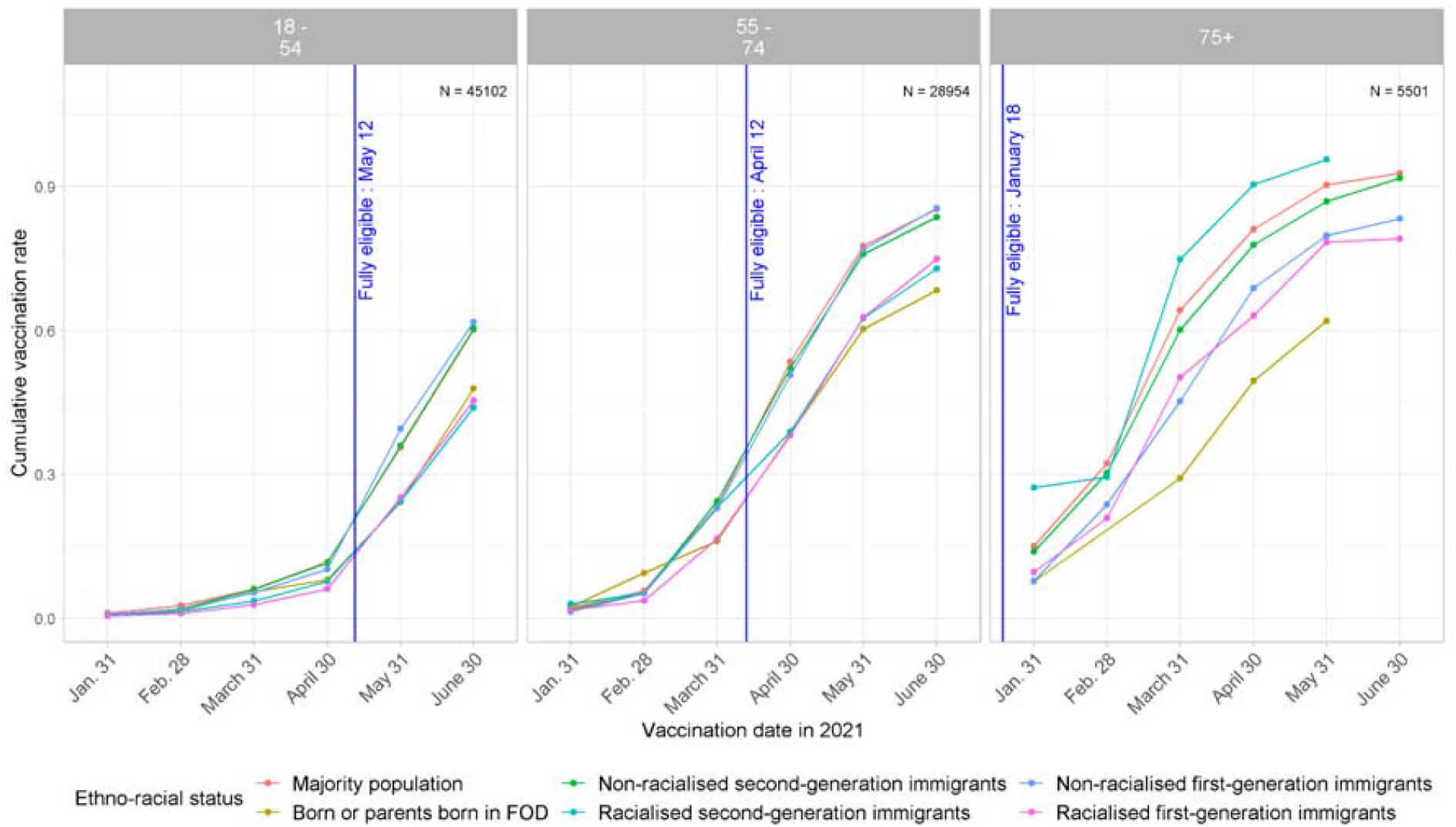
Trends over time in vaccination cumulative incidence rates by age, according to ethno-racial status. EpiCov study, 3rd wave, July 2021.

The mistrust of the government to manage the epidemic crisis was much more pronounced than the mistrust of the scientists: in all, 15.7% of the respondents trusted the government completely and 17.3% did not trust them at all, while 36.8% of the respondents trusted scientists completely and only 3.9% did not trust them at all (supplementary Table 1). People at the bottom of the social hierarchy showed much more distrust in the government. The differences were similar although less pronounced for trust in scientists. Vaccination status varied greatly and regularly according to the degree of trust in the government : 85.0% of those who trusted the government completely to manage the epidemic were vaccinated, 80.5% of those who had a fair amount of trust, 65.2% for those who had little trust and 51.1% of those who did not trust the government at all. Similarly, only 1.9% of those who trusted the government completely refused to be vaccinated, compared to 25.0% of those who did not trust the government at all. The differences were even more pronounced concerning trust in scientists to solve the epidemic crisis (Table 1).

Multivariate analysis confirmed that the social profiles of those not vaccinated, split between those who reported their intention to be vaccinated, those who did not yet know, and those who did not want to be vaccinated, appeared to differ from those in the vaccinated group (Table 2). They were younger, less educated, had lower incomes and more often belonged to a racialised minority than vaccinated people in all three sub-groups, especially those who refused to be vaccinated. Multivariate analysis also showed that people’s mistrust in the government and scientists were the factors most strongly associated with refusing to be vaccinated, compared to vaccinated people with an OR of 8.91 (95%CI: 7.17 to 11.01) for complete mistrust of the government and an OR of 9.05 (7.69 to 10.7) for complete mistrust in scientists (Table 2). Except for age, these factors were also the most discriminating among people who intended to get vaccinated and to a lower extent among people who did not know yet what to do (Table 2).

**Table 2:** Factors associated with vaccination status (multinomial regression, reference = being vaccinated). EpiCov study 3rd wave, July 2021.

|  | Intends to get vaccinated |  | Does not know yet |  | Refuses to get vaccinated |  |
| --- | --- | --- | --- | --- | --- | --- |
|  | OR | 95% CI | OR | 95% CI | OR | 95% CI |
| **Sex** |  |  |  |  |  |  |
| Men | — | — | — | — | — | — |
| Women | 0.85 | 0.80, 0.90 | 1.02 | 0.96, 1.09 | 1.19 | 1.10, 1.28 |
| **Age** |  |  |  |  |  |  |
| 18 - 24 | — | — | — | — | — | — |
| 25 - 34 | 1.20 | 1.06, 1.36 | 1.34 | 1.18, 1.52 | 1.39 | 1.20, 1.60 |
| 35 - 44 | 0.83 | 0.73, 0.93 | 0.99 | 0.87, 1.11 | 0.90 | 0.78, 1.04 |
| 45 - 54 | 0.54 | 0.48, 0.61 | 0.62 | 0.55, 0.71 | 0.58 | 0.50, 0.67 |
| 55 - 64 | 0.33 | 0.29, 0.38 | 0.47 | 0.40, 0.54 | 0.43 | 0.37, 0.51 |
| 65 - 74 | 0.20 | 0.17, 0.24 | 0.26 | 0.22, 0.31 | 0.24 | 0.20, 0.29 |
| 75 - 84 | 0.10 | 0.07, 0.13 | 0.19 | 0.15, 0.25 | 0.20 | 0.14, 0.27 |
| 85+ | 0.16 | 0.10, 0.25 | 0.20 | 0.13, 0.33 | 0.47 | 0.31, 0.72 |
| **Formal education** |  |  |  |  |  |  |
| High school +5 or more years | — | — | — | — | — | — |
| High school +2 to 4 years | 1.08 | 0.97, 1.19 | 1.20 | 1.08, 1.33 | 1.15 | 1.01, 1.30 |
| High school | 1.31 | 1.17, 1.46 | 1.27 | 1.13, 1.44 | 1.39 | 1.21, 1.61 |
| Vocational secondary | 1.32 | 1.16, 1.50 | 1.39 | 1.21, 1.58 | 1.32 | 1.13, 1.54 |
| Primary education | 1.29 | 1.10, 1.50 | 1.36 | 1.15, 1.59 | 1.25 | 1.03, 1.51 |
| No diploma | 1.32 | 1.11, 1.58 | 1.59 | 1.34, 1.89 | 1.39 | 1.13, 1.71 |
| **Standard of living (in deciles)** |  |  |  |  |  |  |
| D10 | — | — | — | — | — | — |
| D8-D9 | 1.11 | 1.00, 1.24 | 1.40 | 1.24, 1.58 | 1.28 | 1.11, 1.48 |
| D6-D7 | 1.26 | 1.12, 1.40 | 1.51 | 1.33, 1.71 | 1.66 | 1.43, 1.92 |
| D4-D5 | 1.53 | 1.36, 1.71 | 1.76 | 1.55, 2.00 | 1.69 | 1.45, 1.97 |
| D2-D3 | 1.70 | 1.51, 1.93 | 2.13 | 1.86, 2.43 | 2.12 | 1.81, 2.48 |
| D1 | 1.96 | 1.71, 2.25 | 2.37 | 2.05, 2.74 | 2.47 | 2.08, 2.93 |
| **Ethno-racial status** |  |  |  |  |  |  |
| Mainstream population | — | — | — | — | — | — |
| Born or parents born in FOD | 1.46 | 1.16, 1.85 | 1.55 | 1.23, 1.96 | 1.49 | 1.13, 1.96 |
| Non-racialised second-generation immigrants | 1.11 | 0.98, 1.26 | 1.14 | 1.00, 1.30 | 1.07 | 0.92, 1.25 |
| Racialised second-generation immigrants | 1.31 | 1.14, 1.51 | 1.95 | 1.71, 2.23 | 1.57 | 1.33, 1.87 |
| Non-racialised first-generation immigrants | 0.94 | 0.78, 1.13 | 0.94 | 0.77, 1.14 | 1.21 | 0.98, 1.50 |
| Racialised first-generation immigrants | 1.70 | 1.47, 1.96 | 1.93 | 1.65, 2.24 | 1.58 | 1.29, 1.94 |
| **Trust in the government** |  |  |  |  |  |  |
| Complete trust | — | — | — | — | — | — |
| Fair amount of trust | 1.18 | 1.06, 1.31 | 1.21 | 1.05, 1.40 | 1.00 | 0.80, 1.25 |
| Little trust | 1.57 | 1.40, 1.75 | 2.48 | 2.15, 2.87 | 3.17 | 2.55, 3.94 |
| Not trust at all | 1.79 | 1.58, 2.03 | 3.12 | 2.68, 3.63 | 8.86 | 7.13, 11.0 |
| **Trust in the scientists** |  |  |  |  |  |  |
| Complete trust | — | — | — | — | — | — |
| Fair amount of trust | 1.17 | 1.09, 1.25 | 2.25 | 2.07, 2.45 | 2.87 | 2.55, 3.22 |
| Little trust | 1.34 | 1.18, 1.52 | 4.08 | 3.63, 4.58 | 8.62 | 7.53, 9.87 |
| Not trust at all | 1.22 | 0.99, 1.51 | 3.19 | 2.67, 3.80 | 9.07 | 7.71, 10.7 |

The data also showed that the richer the people, the more the effects of trust in the government on the decision not to refuse to get vaccinated (Figure 2 and supplementary Table 2a). Compared to the 10% richest people who trust the government, the 10% poorest people who also did reached an OR of 4.44 (3.13 to 6.31) for the decision to refuse to get vaccinated,, the 10% poorest people who did not trust the government reached an OR of 16.2 (11.9 to 22.0) (Figure 2 and supplementary Table 2a). Similar but less marked differences were found according to formal education. Finally, the effect of trust in the government on decreasing refusal to get vaccinated was less pronounced among the racialised first and second generations compared to the mainstream population.

**Figure 2:**
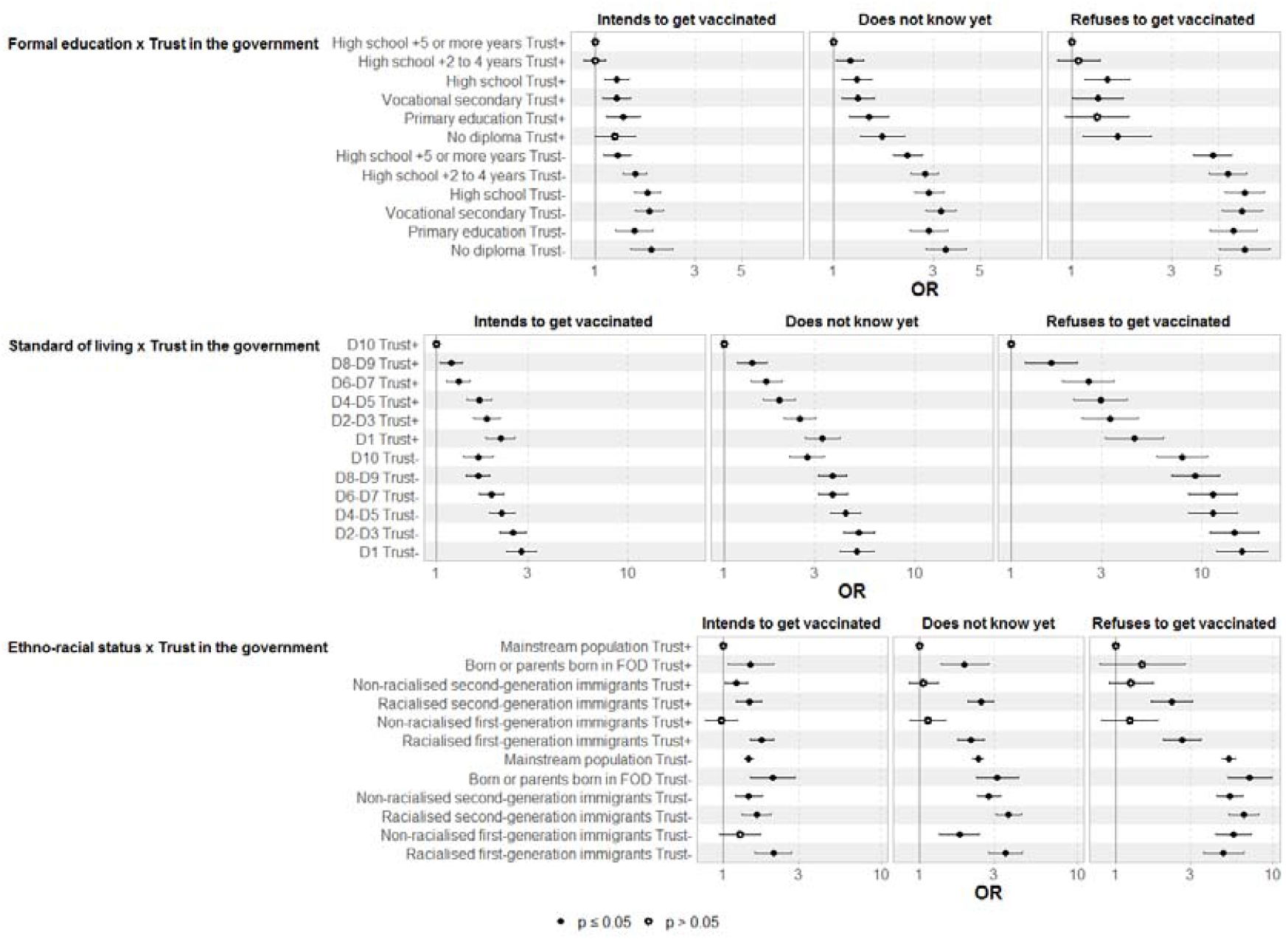
Interaction between trust in the government (yes or no) and *(i)* diploma, *(ii)* standard of living and *(iii)* ethno racial status and vaccination status. Multinomial regression (ref = being vaccinated). EpiCov study, 3rd wave, July 2021. *Also adjusted for age, sex, social class, healthcare worker, cohabitation status, has children, population size of municipality, priority neighbourhood, perceived health status, Covid-19 comorbidities, knows someone who has had a severe form of Covid-19, Covid-19 risk perception, positive test in last 6 months, date of response to questionnaire and trust in scientists*

The results were similar but to a lesser extent for mistrust in scientists (Supplementary Table 2b).

## DISCUSSION

EpiCov is among the largest national socio-epidemiological cohort surveys to be conducted on a random sample of the population, simultaneously taking into account gender, class and ethno-racial status, health data, and mistrust of the government and of scientists to analyse social inequalities in vaccination.

We found marked social and ethnoracial inequalities in vaccination, in a context of free access to vaccination and at a time when anti-Covid certifications had not yet been made restrictive. The least educated, those with the lowest incomes, and racialised minorities were less likely to have been vaccinated and these differences were maintained or increased over time. People’s mistrust in the government and scientists to manage the health crisis remained the factor most strongly associated with refusing to get vaccinated. The impact of trust on not refusing to be vaccinated was even more marked among people at the top of the social hierarchy, thus reinforcing social inequalities in vaccination.

With regard to the social barriers to access to vaccination, we should first of all note that the lower vaccination rates among younger people are likely to be related to a shorter access period. Indeed, data was collected in July 2021, at a time when Covid-19 vaccines were available in France for any individuals aged 18 and over as of May 12th 2021 (12th April for people over 55, and 18th January 2021 for people over 75). Secondly, it is interesting to note that social differences in vaccination practices overlapped with the social distribution of vaccine reluctance observed in France eight months earlier, except for gender differences.[13] Indeed, women were no less likely to be vaccinated than men, although they were more reluctant to get vaccinated in France, as in many countries, before the vaccine was made available to all.[31] Faced with the reality/possibility of prevention, it is as if their gendered reflexes as guardians of the family’s health came into play.[32] Our survey also showed that those with lower levels of education and those belonging to the working class were less likely to be vaccinated, as found in a British survey covering the first 100 days of the vaccine rollout.[7] This could relate to the fact that members of the working classes have a perception of their body and their health that is more distant from medical diagnoses and recommendations than the upper class.[33] Racialised minorities, who had greater reluctance towards receiving a Covid-19 vaccine in France and in many countries[13,31] also appeared to be less likely to be vaccinated, as found in British and US surveys.[6,8,20,34] Numerous studies have shown that racialised minorities[24,35] have less confidence in the healthcare system and in caregivers than the mainstream population.[36,37] This mistrust results in particular from discrimination and mistreatment to which these populations have been exposed when resorting to the public health system.[38,39] A recent study among students in London showed that experiences of racial discrimination increased the likelihood of subsequent Covid-19 vaccine refusal nearly 4-fold.[8]. Barriers other than experiences of discrimination should also be considered, such as the lack of health insurance coverage in countries where vaccination is not free.[40]. In this respect, it is surprising to note that here significant differences were recorded according to income level in the multivariate model, despite vaccination being free in France. While the poorest people have the same tendency as others to comply with the use of masks in France,[41] they are less likely to be vaccinated. One might think that the level of income here reflects above all a certain degree of social integration. The exclusion of the poorest part of the population from the social contract could lead to a diminished sensitivity towards the national solidarity dimension of vaccination, strongly emphasised in the public discourse on prevention in France. The low rates of vaccination among the most deprived, also found in a US survey [9], probably also relates to the fact that they generally have poor access to healthcare than others for given needs.[12]

Our results underline the need to develop outreach strategies targeting the poorest, the least educated people, and the racialised minorities, as recommended by Hanif back in 2020.[42] However, given the preponderant place of vaccine refusal due to a mistrust of the government’s and scientists’ attempts to curb the spread of the coronavirus, the characteristics of the messenger in vaccination campaigns should also be considered. Several studies in the US have shown that non-uptake of vaccination is higher in counties where conservative votes are higher[15,16]. Another study compared the relationship between government trust and vaccination coverage in 177 countries, using pre-pandemic trust scores [43]. However, it is possible to trust politicians to manage the health crisis, even if they do not represent one’s own political ideas. Furthermore, in a context where citizens position themselves less and less within a right-left divide or in the political institutions [44], especially in France [45], it seems more relevant to consider the link between trust in the government and individual decision-making about vaccination. Therefore, the political dimension of vaccination resistance needs to be measured, even for people who have no political opinions or refuse to express them. We found that mistrust in the government and scientists to curb the spread of the epidemic was the strongest predictor for not being vaccinated. Nevertheless the protective effects of trust were less pronounced for people at the lower end of the social ladder and for racialised minorities, with the reinforcement of social inequalities in vaccination as a consequence. It thus seems preferable for the preventive discourse to come from health agencies in close collaboration with community organisations and social workers [46] without political interference. People’s mistrust of scientists, also found in a US survey studying racial differences in vaccination uptake[20], could reflect a strong connivance, in France, between the government and the scientific council. It could also reflect doubts arising from the contradictory injunctions that have been made in the media. Finally, suspicions of scientists colluding with big pharmaceutical companies, following comments made on social media, could also contribute to explaining this mistrust.[47]

Our analysis nevertheless has some limitations. First, as any national population-based survey, the present study failed to capture highly vulnerable groups such as undocumented migrants and homeless people, who were particularly affected by the pandemic.[34]

Secondly, our analysis was based on a survey conducted in July 2021. Until reaching a plateau in October 2021,[49] vaccination rates continued to rise particularly in connection with the mandatory anti-Covid-19 certification (requiring proof of vaccination, recent negative test, or proof of recovery to access specific places such as restaurants, theatres, trains, planes, etc.) introduced on July 21st 2021, which increased vaccine uptake.[2,50] Considering that the least privileged social groups are less impacted by the anti-Covid-19 certification, since they are not likely to routinely access such places, we could hypothesise that the social inequalities observed are still present today, even if their magnitude is less prominent. In addition, it was interesting to study the social inequalities in vaccination practises before the introduction of the mandatory anti-Covid-19 certification to be able to evaluate its effectiveness afterwards.

The highly structuring effect of mistrust in the government and scientists remains to be understood in greater detail. The role of the social networks and the contradictory information on Covid-19 vaccination [20] is particularly difficult to grasp in a quantitative survey, and has not been taken into account.

It should also be emphasised that the spread of the Omicron variant, which has led to a further outbreak of the epidemic in France and in many countries with high vaccination rates, raises questions for many people about the effectiveness of vaccination. New strains, the requirement for boosters, the uncertainty of a possible herd immunity,[51] and the complexity of the scientific and political discourse on Covid-19 vaccines could prompt concerns that groups of people in the population who are more distant from health literacy may no longer embrace the Covid-19 vaccine.

Finally, the issue of social inequalities in vaccination practises is all the more important because the social groups that are the least vaccinated are also those most at risk of contracting Covid-19. Our analyses show that a top-down conception of preventive policies comes up against the social logics that structure the decision to get vaccinated. There is an urgent need to depoliticise vaccination strategies, and to develop outreach programmes for the most socially disadvantaged groups but also “culturally competent” vaccination campaigns [42] conceived with people from different social and racial backgrounds to enable them to understand the scientific and public health challenges of vaccination and make fully informed choices.

## SUMMARY BOXES

### Section 1: What is already known on this topic

Some studies in the UK and in the US have shown that the most socially disadvantaged and racialised social groups are the least vaccinated, and others have shown that the conservative political vote was associated with lower rates of vaccination in the US

### Section 2: What this study adds

We found social and ethnoracial inequalities in vaccination practises, which result from social barriers to engaging in prevention practices. But above all, people’s mistrust in the government and scientists was the factor most strongly associated with refusing to get vaccinated. Nevertheless the effects of trust on not refusing to get vaccinated were less pronounced for people at the lower end of the social ladder and for those who belong to racialised minorities, leading to the reinforcement of social inequalities in vaccination. Our results show the need to develop outreach strategies with no interference of politics, delegated to key-players able to design targeted preventive messages conceived with people from different social and racial backgrounds to enable people to make fully informed choices.

## Data Availability

Anonymous aggregated data for the first round are available online. The EpiCov dataset is available for research purpose concerning the first round, and will be available by March 2022 concerning the second round for research purpose on CASD (https://www.casd.eu/), after submission to approval of French Ethics and Regulatory Committee procedure (Comite du Secret Statistique, CESREES and CNIL).  Access to anonymized individual data underlying the findings may be available before the planned period, on request to the corresponding author, to be submitted to approval of the ethics and reglementary Committee for researchers who meet the criteria for access to data.

## ACKNOWLEDGMENTS

The authors warmly thank all the volunteers of the EpiCov cohort.

We thank the DREES and INSEE teams for the creation of the sample, the calculation of the survey weights and the logistics of the survey.

We thank the staff of the IPSOS team that have worked with dedication and engagement to collect and manage the data used for this study

We thank Frédéric Robergeau, Inserm, responsible for the management of the database and Carmen Calandra, Inserm, project manager.

## DATA SHARING STATEMENT

Anonymous aggregated data for the first round are available online. The EpiCov dataset is available for research purpose concerning the first round, and will be available by March 2022 concerning the second round for research purpose on CASD (https://www.casd.eu/), after submission to approval of French Ethics and Regulatory Committee procedure (Comité du Secret Statistique, CESREES and CNIL). Access to anonymized individual data underlying the findings may be available before the planned period, on request to the corresponding author, to be submitted to approval of the ethics and reglementary Committee for researchers who meet the criteria for access to data.

## AUTHORS AND CONTRIBUTORS

Bajos and Spire had full access to all the data in the study and took responsibility for the integrity of the data and the accuracy of the data analysis.

Study concept and design: Bajos, Spire

Data acquisition: IPSOS.

Data analysis and interpretation: Bajos, Spire, Franck, Silberzan

Drafting of the manuscript: Bajos, Spire, Franck, Silberzan

Critical revision of the manuscript for important intellectual content: All authors.

Statistical analysis: Sireyjol

Study supervision: Bajos and Warszawski

## COPYRIGHT

I, the Submitting Author has the right to grant and does grant on behalf of all authors of the Work (as defined in the author licence), an exclusive licence and/or a non-exclusive licence for contributions from authors who are: i) UK Crown employees; ii) where BMJ has agreed a CC-BY licence shall apply, and/or iii) in accordance with the terms applicable for US Federal Government officers or employees acting as part of their official duties; on a worldwide, perpetual, irrevocable, royalty-free basis to BMJ Publishing Group Ltd (“BMJ”) its licensees.

The Submitting Author accepts and understands that any supply made under these terms is made by BMJ to the Submitting Author unless you are acting as an employee on behalf of your employer or a postgraduate student of an affiliated institution which is paying any applicable article publishing charge (“APC”) for Open Access articles. Where the Submitting Author wishes to make the Work available on an Open Access basis (and intends to pay the relevant APC), the terms of reuse of such Open Access shall be governed by a Creative Commons licence – details of these licences and which licence will apply to this Work are set out in our licence referred to above.

## TRANSPARENCY STATEMENT

The lead author affirms that this manuscript is an honest, accurate, and transparent account of the study being reported; that no important aspects of the study have been omitted; and that any discrepancies from the study as planned (and, if relevant, registered) have been explained.

## ROLE OF THE STUDY SPONSORS AND STATEMENT OF INDEPENDENCE OF RESEARCHERS FROM FUNDERS

The funders facilitated data acquisition but had no role in the design, analysis, interpretation, or writing.

## ETHICS APPROVAL

The survey was approved by the CNIL (French independent administrative authority responsible for data protection) on 25 April 2020 (ref: MLD/MFI/AR205138) and by the ‘Comité de protection des personnes’ (French equivalent of the Research Ethics Committee) on 24 April. The survey also obtained an agreement from the ‘Comité du Label de la statistique publique’, proving its adequacy to statistical quality standards.

## PATIENT AND PUBLIC STATEMENT

Patients or the public were not involved in the design, or conduct, or reporting, or dissemination plans of our research.

