## Supplemental Table for "When mistrust in the government and scientists reinforce social inequalities in vaccination against Covid-19"

**Supplementary Table 1: Mistrust of the government and scientists according to social characteristics**

|  | **Trust in the government** | | | | **Trust in scientists** | | | |
| --- | --- | --- | --- | --- | --- | --- | --- | --- |
|  | **Complete trust** | **Fair amount of trust** | **Little trust** | **No trust at all** | **Complete trust** | **Fair amount of trust** | **Little trust** | **No trust at all** |
| **Total** | 15.7 | 41.3 | 24.9 | 17.3 | 36.8 | 51.6 | 6.9 | 3.9 |
| **Sex** |  |  |  |  |  |  |  |  |
| Men | 17.2 | 41.0 | 23.3 | 17.9 | 39.1 | 49.7 | 6.7 | 4.1 |
| Women | 14.3 | 41.5 | 26.4 | 16.7 | 34.8 | 53.5 | 7.2 | 3.7 |
| **Age** |  |  |  |  |  |  |  |  |
| 18 - 24 | 10.2 | 39.5 | 30.8 | 19.1 | 32.8 | 53.0 | 9.1 | 4.7 |
| 25 - 34 | 8.7 | 37.1 | 31.7 | 21.9 | 27.6 | 55.3 | 11.0 | 5.7 |
| 35 - 44 | 12.6 | 40.2 | 27.4 | 19.3 | 29.1 | 55.6 | 9.8 | 4.9 |
| 45 - 54 | 14.8 | 41.8 | 24.5 | 18.1 | 33.9 | 54.2 | 6.9 | 4.5 |
| 55 - 64 | 16.5 | 43.1 | 23.4 | 16.2 | 39.2 | 51.5 | 5.2 | 3.4 |
| 65 - 74 | 22.4 | 42.1 | 20.5 | 14.3 | 47.0 | 46.1 | 3.9 | 2.3 |
| 75 - 84 | 25.5 | 43.0 | 17.3 | 12.2 | 50.8 | 43.4 | 3.0 | 1.6 |
| 85+ | 23.1 | 50.1 | 15.1 | 9.6 | 46.5 | 47.8 | 2.7 | 1.8 |
| **Social class** |  |  |  |  |  |  |  |  |
| Self-employed and entrepreneurs | 21.0 | 41.6 | 20.9 | 15.5 | 39.3 | 49.2 | 7.1 | 3.8 |
| Senior executive professionals | 17.1 | 49.2 | 22.2 | 11.2 | 42.7 | 51.3 | 4.5 | 1.3 |
| Middle executive professionals | 13.0 | 43.3 | 27.3 | 15.8 | 35.8 | 54.2 | 6.8 | 2.8 |
| Employees | 14.6 | 38.9 | 26.2 | 19.2 | 34.0 | 52.7 | 7.9 | 4.4 |
| Manual workers | 16.9 | 34.7 | 24.2 | 23.2 | 35.7 | 48.7 | 8.3 | 6.4 |
| Students | 10.8 | 43.5 | 29.2 | 16.0 | 36.6 | 51.6 | 7.6 | 3.7 |
| Never worked | 23.7 | 39.1 | 20.8 | 15.2 | 36.3 | 50.7 | 5.6 | 6.2 |
| Farmers | 20.3 | 41.3 | 21.1 | 14.7 | 36.1 | 52.9 | 5.5 | 3.7 |
| Missing | 164 | 322 | 218 | 222 | 344 | 438 | 80 | 65 |
| **Formal education** |  |  |  |  |  |  |  |  |
| No diploma | 23.2 | 34.5 | 18.7 | 21.5 | 40.4 | 44.6 | 6.3 | 7.0 |
| Primary education | 19.8 | 41.2 | 21.5 | 15.9 | 43.2 | 47.1 | 5.5 | 3.0 |
| Vocational secondary | 15.8 | 37.2 | 25.0 | 21.3 | 35.5 | 50.9 | 7.2 | 5.7 |
| High school | 12.8 | 40.4 | 27.5 | 18.8 | 33.1 | 53.5 | 8.7 | 4.2 |
| High school +2 to 4 years | 12.4 | 44.8 | 28.0 | 14.4 | 33.7 | 56.3 | 7.3 | 2.4 |
| High school +5 or more years | 15.2 | 49.9 | 24.2 | 10.4 | 41.1 | 52.6 | 5.0 | 1.1 |
| **Standard of living (in deciles)** |  |  |  |  |  |  |  |  |
| D1 | 18.8 | 34.7 | 23.6 | 22.0 | 35.9 | 47.0 | 9.3 | 7.0 |
| D2-D3 | 16.3 | 34.8 | 26.0 | 21.9 | 34.5 | 50.0 | 8.8 | 5.7 |
| D4-D5 | 13.8 | 39.0 | 26.0 | 20.1 | 34.9 | 51.8 | 7.9 | 4.4 |
| D6-D7 | 14.8 | 41.7 | 26.2 | 16.4 | 36.5 | 53.2 | 6.4 | 3.4 |
| D8-D9 | 15.3 | 47.1 | 24.3 | 12.8 | 39.3 | 53.1 | 5.3 | 2.0 |
| D10 | 19.3 | 50.7 | 20.1 | 9.5 | 43.3 | 51.2 | 3.8 | 1.3 |
| Missing | 258 | 834 | 538 | 315 | 678 | 1066 | 142 | 67 |
| **Ethno-racial status** |  |  |  |  |  |  |  |  |
| Mainstream population | 14.3 | 41.7 | 25.7 | 17.5 | 36.4 | 52.7 | 6.8 | 3.6 |
| Born or parents born in FOD | 12.1 | 36.2 | 29.3 | 21.3 | 27.6 | 50.7 | 12.2 | 8.9 |
| Non-Racialised second-generation immigrants | 14.4 | 43.1 | 25.5 | 16.3 | 37.9 | 51.6 | 6.5 | 3.2 |
| Racialised second-generation immigrants | 13.8 | 38.6 | 26.9 | 20.2 | 32.1 | 51.1 | 9.5 | 6.8 |
| Non-Racialised first-generation immigrants | 20.7 | 43.9 | 22.1 | 12.1 | 41.9 | 48.4 | 6.4 | 2.9 |
| Racialised first-generation immigrants | 33.0 | 39.3 | 14.4 | 11.8 | 45.0 | 42.1 | 6.7 | 4.4 |
| Missing | 174 | 449 | 322 | 267 | 416 | 641 | 86 | 72 |

*Data is presented in percentages except for missing values where only numbers are reported. Among men, 17.2% had complete trust in the government at the time of the survey. Individuals who answered “I don’t know” for mistrut variables are not presented*

**Supplementary Table 2a: Interaction between trust in the government (yes or no) and *(i)* diploma, *(ii)* standard of living and *(iii)* ethno-racial status and vaccination status. Multinomial regression (ref = being vaccinated). EpiCov study 3rd wave, July 2021.**

|  | **Intends to get vaccinated** | | **Does not know yet** | | **Refuses to get vaccinated** | |
| --- | --- | --- | --- | --- | --- | --- |
| **Characteristic** | **OR** | **95% CI** | **OR** | **95% CI** | **OR** | **95% CI** |
| **Formal education x Trust in the government** |  |  |  |  |  |  |
| High school +5 or more years Trust+ | — | — | — | — | — | — |
| High school +2 to 4 years Trust+ | 1.00 | 0.88, 1.13 | 1.20 | 1.04, 1.39 | 1.08 | 0.86, 1.37 |
| High school Trust+ | 1.27 | 1.11, 1.45 | 1.29 | 1.09, 1.52 | 1.48 | 1.16, 1.89 |
| Vocational secondary Trust+ | 1.27 | 1.09, 1.48 | 1.31 | 1.10, 1.57 | 1.34 | 1.02, 1.76 |
| Primary education Trust+ | 1.36 | 1.14, 1.64 | 1.48 | 1.19, 1.84 | 1.32 | 0.94, 1.87 |
| No diploma Trust+ | 1.24 | 1.00, 1.55 | 1.71 | 1.35, 2.17 | 1.65 | 1.14, 2.41 |
| High school +5 or more years Trust- | 1.28 | 1.10, 1.48 | 2.26 | 1.93, 2.65 | 4.70 | 3.80, 5.82 |
| High school +2 to 4 years Trust- | 1.55 | 1.36, 1.76 | 2.73 | 2.36, 3.15 | 5.56 | 4.53, 6.82 |
| High school Trust- | 1.78 | 1.55, 2.05 | 2.86 | 2.45, 3.35 | 6.69 | 5.41, 8.28 |
| Vocational secondary Trust- | 1.82 | 1.56, 2.12 | 3.25 | 2.75, 3.83 | 6.51 | 5.21, 8.14 |
| Primary education Trust- | 1.54 | 1.26, 1.89 | 2.85 | 2.32, 3.49 | 5.91 | 4.57, 7.63 |
| No diploma Trust- | 1.86 | 1.47, 2.34 | 3.44 | 2.75, 4.30 | 6.69 | 5.09, 8.80 |
| **Standard of living (in deciles) x Trust in the government** |  |  |  |  |  |  |
| D10 Trust+ | — | — | — | — | — | — |
| D8-D9 Trust+ | 1.19 | 1.04, 1.36 | 1.41 | 1.18, 1.68 | 1.63 | 1.19, 2.23 |
| D6-D7 Trust+ | 1.30 | 1.13, 1.50 | 1.68 | 1.40, 2.02 | 2.54 | 1.86, 3.47 |
| D4-D5 Trust+ | 1.68 | 1.45, 1.94 | 1.96 | 1.62, 2.37 | 2.94 | 2.14, 4.06 |
| D2-D3 Trust+ | 1.84 | 1.57, 2.15 | 2.50 | 2.06, 3.05 | 3.30 | 2.36, 4.61 |
| D1 Trust+ | 2.17 | 1.82, 2.58 | 3.29 | 2.67, 4.05 | 4.44 | 3.13, 6.31 |
| D10 Trust- | 1.66 | 1.38, 1.98 | 2.73 | 2.22, 3.36 | 7.87 | 5.80, 10.7 |
| D8-D9 Trust- | 1.65 | 1.43, 1.90 | 3.71 | 3.12, 4.41 | 9.26 | 6.95, 12.3 |
| D6-D7 Trust- | 1.93 | 1.67, 2.23 | 3.72 | 3.12, 4.44 | 11.4 | 8.55, 15.2 |
| D4-D5 Trust- | 2.20 | 1.89, 2.56 | 4.33 | 3.61, 5.18 | 11.4 | 8.48, 15.2 |
| D2-D3 Trust- | 2.51 | 2.14, 2.94 | 5.12 | 4.25, 6.15 | 14.8 | 11.0, 19.9 |
| D1 Trust- | 2.78 | 2.32, 3.33 | 4.96 | 4.04, 6.07 | 16.2 | 11.9, 22.0 |
| **Ethno-racial status x Trust in the government** |  |  |  |  |  |  |
| Mainstream population Trust+ | — | — | — | — | — | — |
| Born or parents born in FOD Trust+ | 1.49 | 1.07, 2.08 | 1.94 | 1.36, 2.75 | 1.47 | 0.79, 2.74 |
| Non-racialised second-generation immigrants Trust+ | 1.21 | 1.03, 1.43 | 1.06 | 0.86, 1.32 | 1.25 | 0.91, 1.72 |
| Racialised second-generation immigrants Trust+ | 1.46 | 1.21, 1.75 | 2.45 | 2.03, 2.95 | 2.28 | 1.68, 3.09 |
| Non-racialised first-generation immigrants Trust+ | 0.97 | 0.77, 1.23 | 1.13 | 0.87, 1.47 | 1.23 | 0.82, 1.84 |
| Racialised first-generation immigrants Trust+ | 1.76 | 1.49, 2.08 | 2.12 | 1.76, 2.56 | 2.64 | 2.01, 3.48 |
| Mainstream population Trust- | 1.45 | 1.36, 1.55 | 2.35 | 2.19, 2.52 | 5.21 | 4.73, 5.75 |
| Born or parents born in FOD Trust- | 2.06 | 1.48, 2.87 | 3.13 | 2.30, 4.26 | 7.09 | 5.15, 9.77 |
| Non-racialised second-generation immigrants Trust- | 1.45 | 1.20, 1.77 | 2.77 | 2.34, 3.28 | 5.34 | 4.41, 6.46 |
| Racialised second-generation immigrants Trust- | 1.62 | 1.31, 2.01 | 3.67 | 3.05, 4.42 | 6.51 | 5.25, 8.08 |
| Non-racialised first-generation immigrants Trust- | 1.27 | 0.94, 1.73 | 1.79 | 1.34, 2.39 | 5.59 | 4.30, 7.28 |
| Racialised first-generation immigrants Trust- | 2.08 | 1.60, 2.71 | 3.51 | 2.75, 4.47 | 4.84 | 3.60, 6.50 |

*Also adjusted for age, sex, social class, healthcare worker, cohabitation status, has children, population size of municipality, priority neighbourhood, perceived health status, Covid-19 comorbidities, knows someone who has had a severe form of Covid-19, Covid-19 risk perception, positive test in last 6 months, date of response to questionnaire and trust in scientists.*

**Supplementary Table 2b: Interaction between trust in the scientists (yes or no) and *(i)* diploma, *(ii)* standard of living and *(iii)* ethno racial status and vaccination status. Multinomial regression (ref = being vaccinated). EpiCov study 3rd wave, July 2021.**

|  | **Intends to get vaccinated** | | **Does not know yet** | | **Refuses to get vaccinated** | |
| --- | --- | --- | --- | --- | --- | --- |
| **Characteristic** | **OR** | **95% CI** | **OR** | **95% CI** | **OR** | **95% CI** |
| **Formal education x Trust in the government** |  |  |  |  |  |  |
| High school +5 or more years Trust+ | — | — | — | — | — | — |
| High school +2 to 4 years Trust+ | 1.08 | 0.98, 1.20 | 1.26 | 1.12, 1.41 | 1.16 | 1.01, 1.34 |
| High school Trust+ | 1.30 | 1.16, 1.46 | 1.37 | 1.20, 1.55 | 1.48 | 1.26, 1.73 |
| Vocational secondary Trust+ | 1.34 | 1.17, 1.52 | 1.49 | 1.30, 1.72 | 1.36 | 1.15, 1.62 |
| Primary education Trust+ | 1.28 | 1.09, 1.50 | 1.45 | 1.22, 1.72 | 1.31 | 1.06, 1.62 |
| No diploma Trust+ | 1.30 | 1.08, 1.56 | 1.82 | 1.51, 2.18 | 1.43 | 1.12, 1.82 |
| High school +5 or more years Trust- | 1.13 | 0.82, 1.55 | 2.73 | 2.15, 3.47 | 4.04 | 3.18, 5.13 |
| High school +2 to 4 years Trust- | 1.27 | 1.03, 1.56 | 2.89 | 2.44, 3.42 | 4.87 | 4.09, 5.80 |
| High school Trust- | 1.61 | 1.32, 1.98 | 2.71 | 2.26, 3.25 | 5.27 | 4.38, 6.34 |
| Vocational secondary Trust- | 1.42 | 1.13, 1.79 | 2.76 | 2.27, 3.35 | 4.99 | 4.10, 6.08 |
| Primary education Trust- | 1.63 | 1.15, 2.32 | 2.79 | 2.07, 3.76 | 4.64 | 3.49, 6.16 |
| No diploma Trust- | 1.72 | 1.18, 2.50 | 2.47 | 1.78, 3.41 | 4.99 | 3.69, 6.75 |
| **Standard of living (in deciles) x Trust in scientists** |  |  |  |  |  |  |
| D10 Trust+ | — | — | — | — | — | — |
| D8-D9 Trust+ | 1.15 | 1.03, 1.29 | 1.40 | 1.23, 1.59 | 1.29 | 1.08, 1.53 |
| D6-D7 Trust+ | 1.32 | 1.17, 1.48 | 1.58 | 1.39, 1.81 | 1.79 | 1.51, 2.13 |
| D4-D5 Trust+ | 1.57 | 1.39, 1.78 | 1.80 | 1.56, 2.07 | 1.92 | 1.60, 2.30 |
| D2-D3 Trust+ | 1.78 | 1.56, 2.02 | 2.23 | 1.93, 2.58 | 2.28 | 1.89, 2.75 |
| D1 Trust+ | 2.02 | 1.75, 2.33 | 2.58 | 2.20, 3.01 | 2.70 | 2.21, 3.30 |
| D10 Trust- | 2.00 | 1.44, 2.78 | 2.92 | 2.17, 3.95 | 5.50 | 4.16, 7.26 |
| D8-D9 Trust- | 1.35 | 1.06, 1.72 | 3.67 | 3.03, 4.45 | 6.26 | 5.10, 7.69 |
| D6-D7 Trust- | 1.30 | 1.02, 1.64 | 2.84 | 2.32, 3.46 | 6.27 | 5.11, 7.70 |
| D4-D5 Trust- | 1.88 | 1.50, 2.35 | 3.81 | 3.12, 4.64 | 6.10 | 4.92, 7.56 |
| D2-D3 Trust- | 1.90 | 1.48, 2.44 | 4.01 | 3.24, 4.96 | 8.05 | 6.44, 10.1 |
| D1 Trust- | 2.33 | 1.76, 3.08 | 3.72 | 2.88, 4.80 | 8.61 | 6.70, 11.1 |
| **Ethno-racial status x Trust in scientists** |  |  |  |  |  |  |
| Mainstream population Trust+ | — | — | — | — | — | — |
| Born or parents born in FOD Trust+ | 1.57 | 1.22, 2.01 | 1.49 | 1.14, 1.95 | 1.70 | 1.22, 2.36 |
| Non-racialised second-generation immigrants Trust+ | 1.14 | 1.00, 1.30 | 1.17 | 1.02, 1.35 | 1.09 | 0.90, 1.31 |
| Racialised second-generation immigrants Trust+ | 1.34 | 1.16, 1.55 | 1.96 | 1.70, 2.27 | 1.58 | 1.29, 1.93 |
| Non-racialised first-generation immigrants Trust+ | 0.93 | 0.77, 1.13 | 0.94 | 0.76, 1.16 | 0.99 | 0.75, 1.30 |
| Racialised first-generation immigrants Trust+ | 1.63 | 1.40, 1.89 | 1.93 | 1.64, 2.27 | 1.90 | 1.52, 2.38 |
| Mainstream population Trust- | 1.19 | 1.06, 1.33 | 2.08 | 1.90, 2.28 | 3.88 | 3.56, 4.23 |
| Born or parents born in FOD Trust- | 1.12 | 0.56, 2.23 | 3.27 | 2.03, 5.28 | 4.59 | 2.82, 7.49 |
| Non-racialised second-generation immigrants Trust- | 1.01 | 0.65, 1.56 | 1.99 | 1.45, 2.74 | 3.66 | 2.77, 4.85 |
| Racialised second-generation immigrants Trust- | 1.35 | 0.88, 2.08 | 3.71 | 2.69, 5.12 | 5.64 | 4.10, 7.78 |
| Non-racialised first-generation immigrants Trust- | 1.20 | 0.62, 2.33 | 1.79 | 1.05, 3.04 | 6.76 | 4.59, 9.96 |
| Racialised first-generation immigrants Trust- | 2.93 | 1.94, 4.42 | 3.81 | 2.61, 5.57 | 3.92 | 2.55, 6.02 |

*Also adjusted for age, sex, social class, healthcare worker, cohabitation status, has children, population size of municipality, priority neighbourhood, perceived health status, Covid-19 comorbidities, knows someone who has had a severe form of Covid-19, Covid-19 risk perception, positive test in last 6 months, date of response to questionnaire and trust in government.*
